# Some Mathematics for the Method of Pooled PCR Test

**DOI:** 10.1101/2021.03.26.21254430

**Authors:** Nobuyuki Otsu, Honorable Researcher of AIST

## Abstract

At the time of the worldwide COVID-19 disaster, the author learned about the pooled (RT-) PCR test from the news. From the common sense of individual tests, the idea of mixing multiple samples seems taboo, however in fact many samples can be tested with a smaller number of tests by the method. As a retired researcher of mathematical engineering, the author was deeply interested in the idea and absorbed in the mathematical formulation and intensive analysis of the method.

Later, he found that the original basic equation was already proposed in the old (1943) treatise [1] and so many related research works have been done and available as materials on the web [2], although many of those seem to be based on qualitative or intuitive analysis. In that sense, some of the analysis here seems to be already known in the field, but some results might be novel, such as boundary conditions, derivation of limit values, estimation of infection rate and adaptive optimization scheme of pool test, strict extension to multi-stage pool test, and explicit derivation of asymptotic approximate solutions of optimal pooling number and achieved efficiency measure, etc.

In any case, he decided to put it together here as a material rather than a formal treatise, hoping that the results here would be useful for deeper mathematical insights into and better understanding of the pool inspection, and also in its actual practice.

## 1. Introduction

To address the coronavirus pandemic, it is desired to expand and boost the infection testing such as (RT-) PCR. It is the basis of infectious disease control to find as many infected people as possible by testing and to quarantine them promptly. ^1)^ However, due to the allowable amount of testing, the test is carried out being delayed or prioritized for (or limited to) those who develop the disease and their close contacts (especially in Japan), which has led to the spread of infection by overlooking potential asymptomatic infected persons with infectivity.

Common sense is that the number of tests for *N* samples is *N* times because it seems necessary to run individual tests, and as *N* increases, the number of tests also increases. However, if more samples can be tested with a smaller number of tests, the test efficiency (throughput) will be greatly improved. In fact, this idea is realized as the pool test method and is performed in the United States, China, etc. with 4 or 5 sample pooling, and it is also a clarification of the huge and rapid PCR test in China.

Recently, numerous related studies have been reported [2], however, such problems remain as how many samples are best to pool, the conditions and application limits. Therefore, in this article, we will mathematically formulate and analyze the efficiency (optimization) of such inspections in intensive ways.

## 2. Pool test method

The proposal of the pool test method is old and seems to date back to Dorfman’s 1943 short treatise [1], which was proposed at the end of World War II as a method for streamlining soldiers’ syphilis testing in the United States. In recent years, it seems that it was also used for the blood donation test, but it has been re-recognized and practiced from the need for PCR test of large samples in this coronavirus disaster. In what follows, we start by mathematically formulating the method.

### 2.1 Definition and formulation

Consider a PCR test of *N* samples (persons), and suppose *s* (0 < *s* < *N*) of these are positive (infected). Then, the probability that one randomly selected sample will be positive is *p* = *s/N* (0 < *p* < 1), and the probability that it will be negative is therefore *q* = 1 − *p*. If the samples from the population are random and the number *N* is large enough, *p* will asymptotically converge to the population infection rate 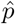 by the law of large numbers. Here, it is assumed that the PCR test is correct (no misjudgment).

Below, a hierarchical (decision tree) test method, or pooled PCR test method is defined.

**S0** : First, *N* samples are randomly divided into *groups* each of which consists of *n* (1 < *n* ≪ *N*) samples (*n* = 5 in China). The last group may consists of less samples. Then, the number of groups is given by ⌈*N/n*⌉ (rounded up integer). Next, the following test procedures are performed for each group.

**S1** : **[Pool Test]** Part (say, half) of each of the *n* samples in the group is taken out, and one sample is made by mixing (pooling) these and tested. If the result is negative, all *n* samples can be judged as negative in this *one* test, and the process is completed. If positive, then go to S2.

**S2** : **[Individual Test]** All *n* samples are further tested individually and judged, and the process is completed. The number of tests in this case is [1 + *n*] times.

Let’s consider to calculate the expected (average) value of the number of tests in this inspection procedure. Since *n* samples in each group are considered random and independent, the probability that a mixed sample will be negative in S1 is *q*^*n*^, and one test of *n* samples is completed. On the other hand, the probability that the mixed sample will be positive is 1 − *q*^*n*^, and in this case, the judgment is completed after *n* more tests. This test is repeated for the number of groups, and therefore the expected value of the total number of tests is given by the following equation (ignoring the fraction in the last group).

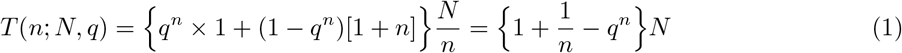

Here, as a measure of test efficiency, consider a value *R* obtained by dividing the number of tests *T* by the number of samples *N*, viz. “tests per samples” (the coefficient on the right side of the above equation).

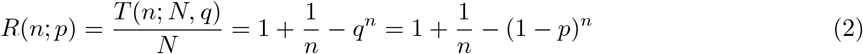

Since *R* = 1 in the case of individual test, if *R* is less than 1, it means that the test has become more efficient. This efficiency measure *R* was already derived in the original paper [1]. It is noted that reciprocal measure 1*/R* is alternatively considered as an efficiency measure indicating “samples per tests”.

### 2.2 Qualitative analysis

The pooling number *n* is 1 < *n* ≪ *N*, but if we consider the boundary conditions of function *R*(*n*; *p*) at both ends, *n* = 1 and *n* = *N*, then we obtain the followings.

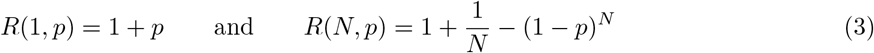

Both cases, however, are reduced to individual test, because pooling is unnecessary in the former case, and pool test results in positive needless to do in the latter case. Actually, it is seen that in both cases *R* converges to 1 (inefficient, same as individual test) for small *p* and large *N*.

While the function *R* is bounded to 1 at both ends *n* = 1 and *N* as stated above, for any *n* in between we obtain the following boundary conditions with respect to *p*.

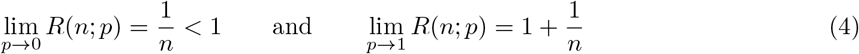

The left equation means that the pooling is effective for smaller *p* and that the continuous function *R* is convex downwards and has the optimal pooling number 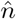 to minimize *R* depending on *p*.

On the other hand, the right equation means that any pooling (*n*) becomes inefficient for larger *p*. This implies that there is a limit value of *p* to make the pool test effective, which will be studied later.

In order to scrutinize the basic equation, Eq. (2), more analytically, let’s consider the partial derivatives by expanding *n* to a real number, too.

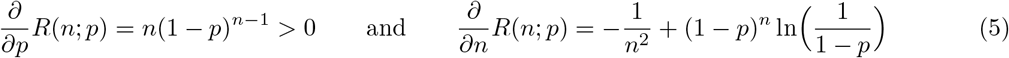

The left equation means that *R* is a monotonically increasing function of *p* for any fixed *n*, which ensures the estimation of *p* from *R* discussed later. The right equation provides the equation for the optimal 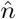, however the complex form makes it difficult to get the explicit solution. Thus, it is left to resort to direct search for the minimum of *R* or to compromise with *ad hoc* setting of *n* to 4 or 5 in actual practices.

In **Fig. 1** the efficiency measure, function *R*(*n*; *p*), is plotted for *n* and some values of *p*. It can be seen that the pool method is rather inefficient for positive rate *p* ≥ 0.3 and the individual test is better. For *p* = 0.1 *n* = 4 is optimal and *R* = 0.594, *n* = 5 is optimal around *p* = 0.05 and *R* = 0.426, and *n* = 6 is optimal for *p* = 0.03 and *R* = 0.334. The smaller the *p*, the larger the optimal 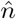, the smaller the *R* value, the wider the valley, viz. the more robust the optimal 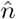 is, and the minimum value of *R* does not change much before and after of 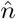. At *p* = 0.003, *n* = 19 is optimal and *R* = 0.108, however the efficiency is almost the same even for *n* = 15 ∼ 23. These results obtained by direct minimum search are listed in **Table 1**.

**Table 1.**
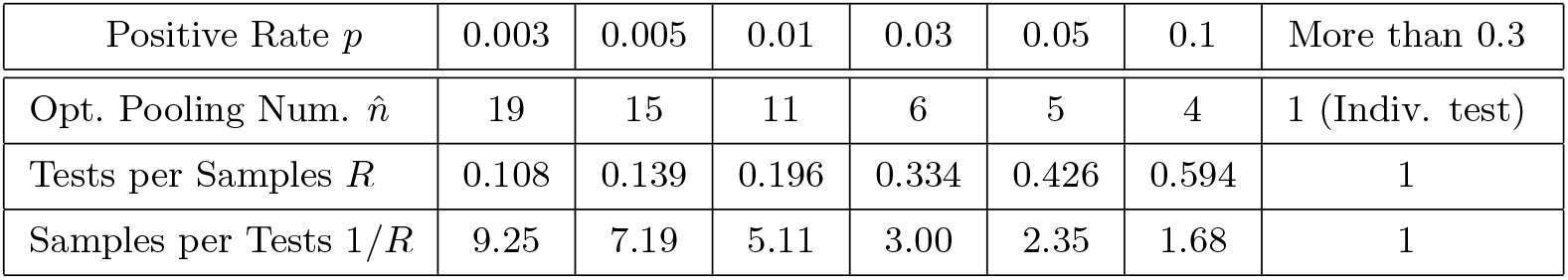
Optimal pooling number 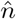 searched and minimum efficiency measure 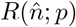 achieved for positive rate *p*

| Positive Rate $p$ | 0.003 | 0.005 | 0.01 | 0.03 | 0.05 | 0.1 | More than 0.3 |
| --- | --- | --- | --- | --- | --- | --- | --- |
| Opt. Pooling Num. $\hat{n}$ | 19 | 15 | 11 | 6 | 5 | 4 | 1 (Indiv. test) |
| Tests per Samples $R$ | 0.108 | 0.139 | 0.196 | 0.334 | 0.426 | 0.594 | 1 |
| Samples per Tests $1/R$ | 9.25 | 7.19 | 5.11 | 3.00 | 2.35 | 1.68 | 1 |

**Fig. 1.**
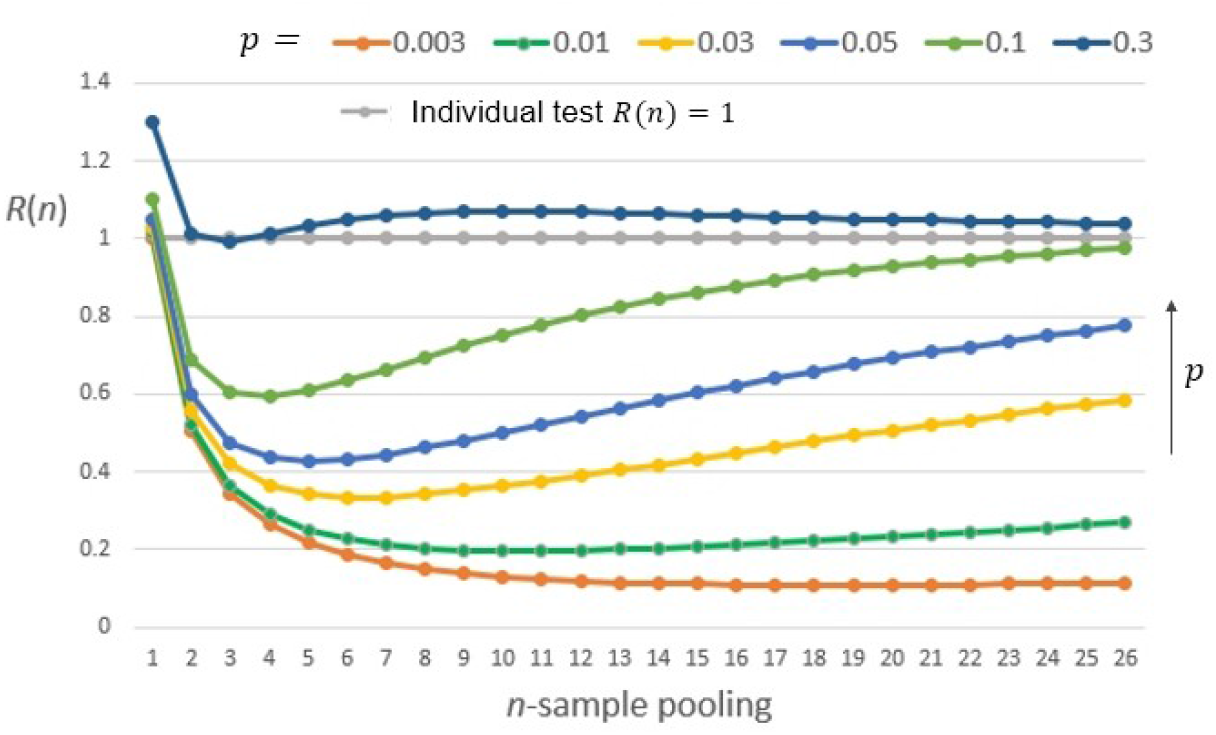
Efficiency measure *R*(*n*; *p*): “tests per samples” for pooling number *n* and positive rate *p*

### 2.3 More analysis including limit value

For understanding above discussions in more details, another analysis of the solution is performed below. Returning to the original Eq. (2), the condition that *R* < 1 (efficient) is given by

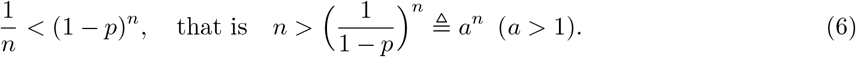

Put the left side as a function 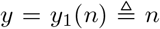 (straight line through the origin) and the right side as a function 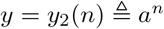 (exponential function), and plot and analyze the graphs for some *p* values.

**Fig. 2** shows the result. The exponential function *y*_2_ is a smooth monotonically increasing function because the base *a* is greater than 1, and for a sufficiently large *n* it is greater than *y*_1_. Also, for *n* = 0, 1, *y*_2_(0) = 1 *> y*_1_(0) = 0, *y*_2_(1) = *a > y*_1_(1) = 1, thus *y*_2_ is larger than *y*_1_. From Eq. (6) the condition for efficiency improvement (*R* < 1) is *y*_1_(*n*) *> y*_2_(*n*), but it is not satisfied at both ends of *n* like this.

**Fig. 2.**
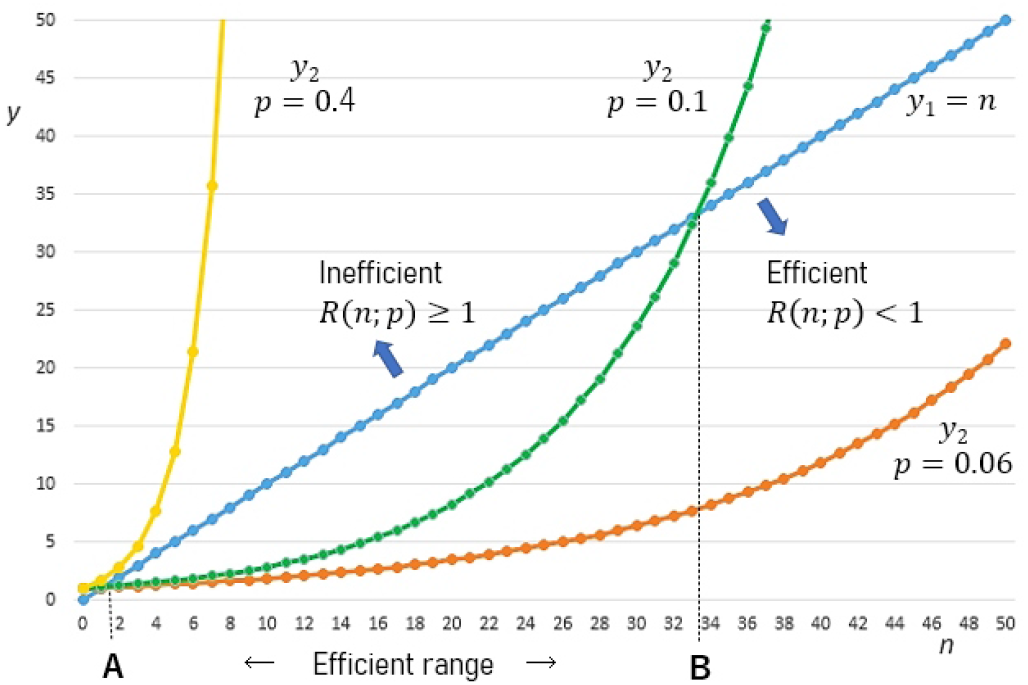
Analysis of *R*(*n*; *p*) using functions *y*_1_ = *n* and *y*_2_ = (1 − *p*)^*−n*^

The larger the positive rate *p*, the larger *a*, and *y*_2_ becomes a rapid increase function and does not intersect with *y*_1_ (the yellow curve for *p* = 0.4 in the figure), but *y*_2_ touches *y*_1_ at *n* = *k* (near 3) with a certain value of *p* (near 0.3), and when *p* becomes smaller than that, *y*_2_ intersects with *y*_1_ at two points *A* and *B*, and the condition of efficiency is satisfied for *n* in between. The effective range [*A, B*] becomes wider as *p* becomes smaller, but as can also be seen from the figure, *A* remains at 1 < *A* < *k* (thus it can be considered *A* = 2 as an integer *n*), while *B* becomes larger. The optimal 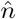 that minimizes *R* in Eq. (2) is the one in the middle of the range [*A, B*].

The condition that the pool method is not valid for any *n*, viz. *R*(*n*; *p*) ≥ 1, is that the value of *p* is greater than the value *p*^∗^ where the function *y*_2_ touches the function *y*_1_ at *n* = *k* and *A* = *B* = *k*. Such limit value *p*^∗^ is found as the solution of the following conditional equations where the tangent of *y*_2_(*n*) at *k* is *y*_1_(*n*).

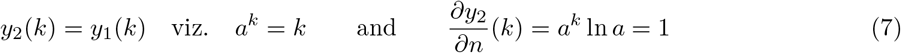

From both equations we have *k* ln *a* = 1, and by taking ln of the right equation we have *k* ln *a*+ln(ln *a*) = 0. By inserting the former into the latter, we have ln(ln *a*) = −1, therefore we obtain the following results.

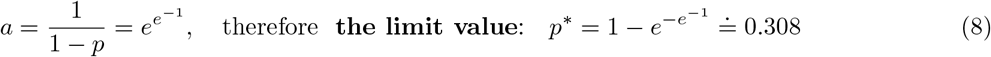

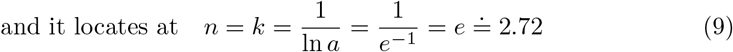

These results analytically explain the graph of *R*(*n*; *p*) when *p* = 0.3 in Fig. 1 obtained by numerical calculation, and the analysis here provides a quantitative analysis for other graphs in Fig. 1.

### 2.4 Estimation of positive rate *p* and adaptive optimization

The above analysis shows that the pool test is ineffective for large positive rate *p* (strictly for *p* ≥ *p*^∗^) and individual test will suffice to do. However, the smaller the positive rate *p*, the more efficient the pool test becomes, and the optimal pooling number 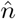 becomes larger. In order to select the optimal 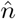, it is first of all necessary to estimate the positive rate *p* of the *N* samples taken from a population. It is, however, not easy, because the positive rate *p* depends on the situation of inspection.

For example, in the case of identifying close contacts in facilities or areas where infection spreads, the number *N* is relatively small, and the samples are not random and highly relevant, so the positive rate *p* is a fairly large value compared with the infection rate 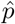 of the larger population. Therefore, individual test or such a pool test as *n* = 4 or 5 seems to be considered rational.

Contrary, in the case of a large mass test to identify potential infected individuals, the samples from the large population are in random sampling and the number *N* becomes quite large. Then, the positive rate *p* = *s/N* as a probability asymptotically converges to the infection rate 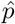 of the population by the law of large numbers. In such cases, 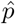 is still a small value, so the enormous number of samples *N* is needed. For example, the average infection rate in Japan is about 0.004 at the moment, so it needs 1000 tests to find 4 positives on average. Therefore, the pool test method is indispensable and effective, and much efficiency improvement can be expected with the optimal pooling number 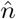 larger than 4 or 5.

In this way, the positive rate *p* depends on the test scale and is basically unknown. While, the overall infection rate 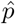 is generally an old statistics (cumulative average) and not necessarily fitted to the current test. Thus, in any case, it is needed to estimate the positive rate *p* of the current test in some way.

Such a method is provided by the original Eq. (2). As has been seen in Eq. (5), the value of efficiency measure *R*(*n*; *p*) with a fixed *n* is monotonically related to *p* (in one-to-one correspondence). Thus, once a pool test on the population is performed with any pre-selected *n* as a trial without knowing *p*, the positive rate *p* can be estimated from the efficiency value (or the average in similar trials) 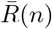 as follows.

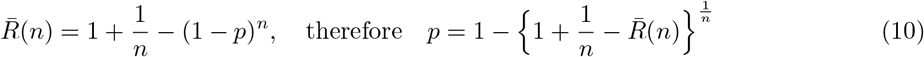

This means that the pool method is well-posed on its inverse problem and *p* (= *s/N*) can be solved inversely and directly from the resultant value 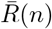 alone without necessitating such additional explicit counting of the number of positives *s* as in the alternative naive estimation method by *p* = *s/N*. Also, 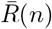 can be calculated from the pool test results at S1, while the naive estimation further requires S2.

By putting this estimated *p* into Eq. (2), the optimal number 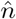 to pool is estimated (searched), and the pool test can be switched to optimized one. This provides an *adaptive optimization scheme* of pool test for preventing the tendency of trial and error or compromise.

### 2.5 First-order approximation of the optimal solution

When the positive rate *p* is 0 < *p* < *p*^∗^, the optimum pooling number 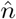 cannot be obtained explicitly. Thus, we have no choice but to resort to the direct search of the minimum value of *R*(*n*; *p*) with changing *n* in steps. However, the approximate solution is obtained by using the McLaughlin (binomial series) expansion of the power function as follows.

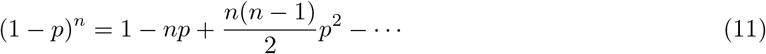

For sufficiently small *p*, the value will be small after the second order, so considering the approximate expression truncated at the first-order term, the following Bernoulli’s inequality holds, and as *p* becomes smaller, it asymptotically converges to the equal sign.

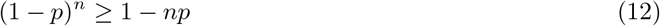

Substituting this into Eq. (2), the upper bound of *R*(*n*; *p*) can be obtained by 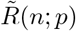 defined as follows.

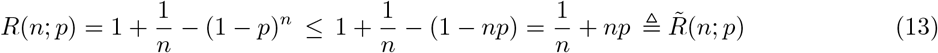

**Fig. 3** shows that the upper bound 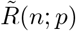 asymptotically converges to *R*(*n*; *p*) as *p* becomes smaller. It should be noticed here that the optimal 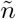 minimizing 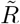 also converges to the optimal 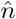 minimizing *R*. This implies that the difficult optimization problem of *R* can be reduced to the approximated one of 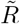.

**Fig. 3.**
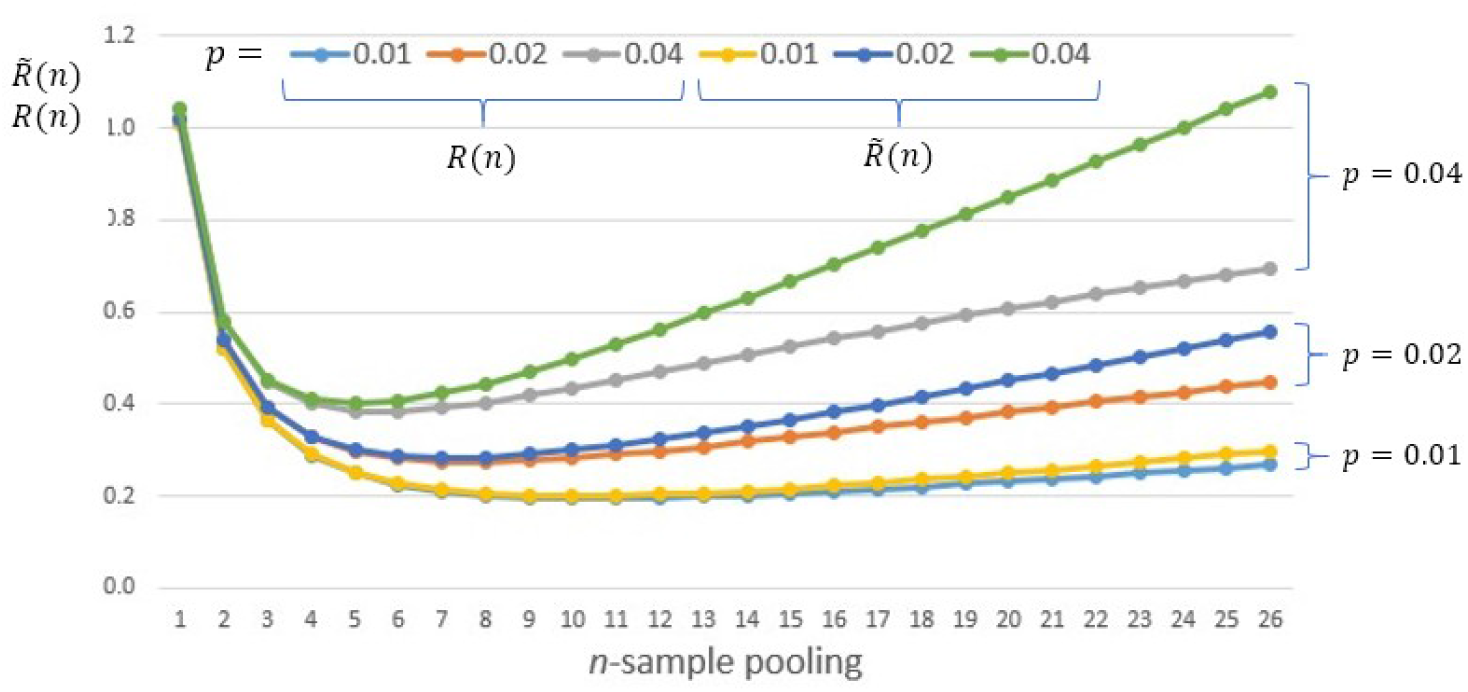
Approximation of *R*(*n*; *p*) by 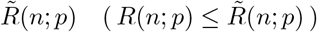 It becomes tighter as positive rate *p* becomes smaller, *p* = 0.04, 0.02, 0.01.

Since 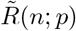 as a function of *n* is convex downward, 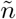, which gives the minimum value, is obtained by differentiating with *n* and setting it as zero.

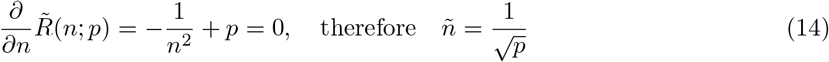

This provides an *explicit* approximate solution of the optimum 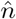 that minimizes *R*. ^2)^ As can be seen in Fig. 3, the approximate solution *ñ* will serve as the lower bound of 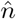, viz. 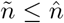 since 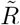 hangs above *R* like strings with the left ends fixed at the same point 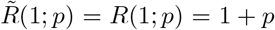. The estimated pooling number *ñ* is generally a real number as 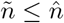 thus considered as rounded up to the nearest integer ⌈*ñ*⌉.

Inserting Eq. (14) to Eq. (13), the approximation (upper bound) of the minimum value of *R* achieved is given by the following explicit simple form.

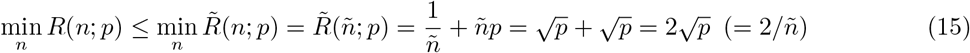

It is noted that these approximate solutions become tightly closer as the positive rate *p* becomes smaller.

From the above analysis, it can be seen that the smaller the positive rate *p*, the larger the approximated 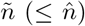 in inverse proportion to 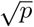, and the smaller the approximated min 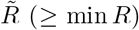 in proportion to 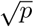, viz. the more efficient the test becomes. Then, it is also seen that the ratio of samples per tests (= 1*/R*) is approximately given by 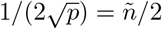. The results of these approximate solutions to Table 1 are shown in **Table 2**. It can be seen that the assumed approximation is better as *p* is smaller.

**Table 2.** Approximate solutions of optimal 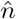 and minimum 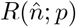 for positive rate *p*: 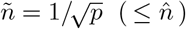 and 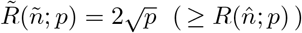 Properly speaking, it states that the estimated optimal pooling number is ⌈*ñ*⌉ and thereby the optimal efficiency measure less (better) than 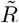 will be achieved.

| Positive Rate $p$ | 0.003 | 0.005 | 0.01 | 0.03 | 0.05 | 0.1 | More than 0.3 |
| --- | --- | --- | --- | --- | --- | --- | --- |
| Opt. Pooling Num. $\tilde{n}$ | 18.26 | 14.14 | 10.00 | 5.77 | 4.47 | 3.16 | 1 (Indiv. test) |
| Tests per Samples $\tilde{R}$ | 0.110 | 0.141 | 0.200 | 0.346 | 0.447 | 0.632 | 1 |
| Samples/Tests $1/\tilde{R}$ | 9.13 | 7.07 | 5.00 | 2.89 | 2.24 | 1.58 | 1 |

In addition, the estimation formula Eq. (10) of the positive rate *p* from the (average) efficiency 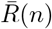 at pre-selected *n* can be easily approximated by the following much simpler formula by using Eq. (13).

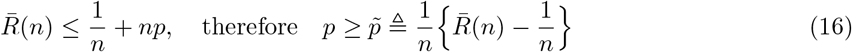

**Table 3** shows the results of the exact estimation formula Eq. (10) obtained in the case of 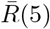(5) and the above approximate estimation formula Eq. (16) at the same *n* = 5. It can be seen that the approximate estimation is sufficiently practical including the inequality sign. Also, it is seen that the smaller the true positive rate *p*, the tighter the approximation 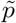, which is preferable in the case of mass inspection.

Using this estimated approximation value 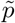, or the exact estimate *p* in Eq. (10), the approximation of the optimum pooling number can be obtained as 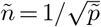 from Eq. (14), and the efficiency measure achieved in that case can be predicted as 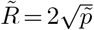 from Eq. (15), which provides the approximated but practical scheme of the adaptive optimization of pool test. For example, if we have 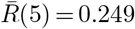 in the pool test with *n* = 5 without knowing *p*, then 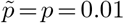 is estimated, and optimal approximate *ñ* = 10 is suggested, so if we switch the test to the case, the efficiency is expected as 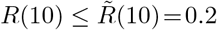, which means that the test is adaptively optimized (improved).

**Table 3.** Estimation of positive rate *p* from (average) efficiency measure 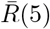 at *n* = 5: 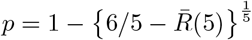 and 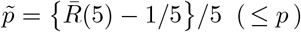

| Positive Rate $p$ | 0.003 | 0.005 | 0.01 | 0.03 | 0.05 | 0.1 |
| --- | --- | --- | --- | --- | --- | --- |
| Ave. Efficiency $\bar{R}(5)$ | 0.215 | 0.225 | 0.249 | 0.341 | 0.426 | 0.610 |
| Exact Estimate $p$ | 0.003 | 0.005 | 0.010 | 0.030 | 0.050 | 0.100 |
| Approximate $\tilde{p}$ | 0.003 | 0.005 | 0.010 | 0.028 | 0.045 | 0.082 |

## 3. Multi-stage pool test method

If *n* in the pool test is large enough (corresponding to the case of small *p*), it is possible to apply an additional pool test for the *n* samples with *m* sample pooling (1 < *m* < *n* ≪ *N*), instead of running individual tests when the group is positive. This idea is natural and has already been referred to in many articles, however in most cases *n* and *m* are independently selected without regard to the total optimization. We consider here the strict formulation of two-stage pool test method and its optimization.

### 3.1 Formulation

In the case of two-stage pool test in the above, the average number of tests *T* for all samples *N* is *recursively* given by the following equation by replacing *n* at [·] in Eq. (1) with *T* (*m*; *n, q*), where *q* = 1 − *p*.

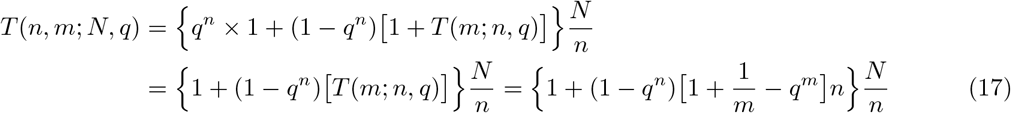

Therefore, the efficiency measure *R* in this case is given by the following equation.

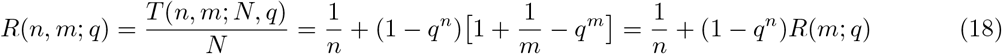

**Table 4** shows the results of the optimal 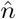 and 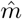 search in this two-stage pool method, corresponding to the one-stage pool method in Table 1. It is seen that further significant efficiency has been achieved.

Interestingly, the optimal number for pooling in the second stage 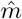 is the same as that in the one-stage pool test, which can be easily seen from Eq. (18) as follows.

**Table 4.**
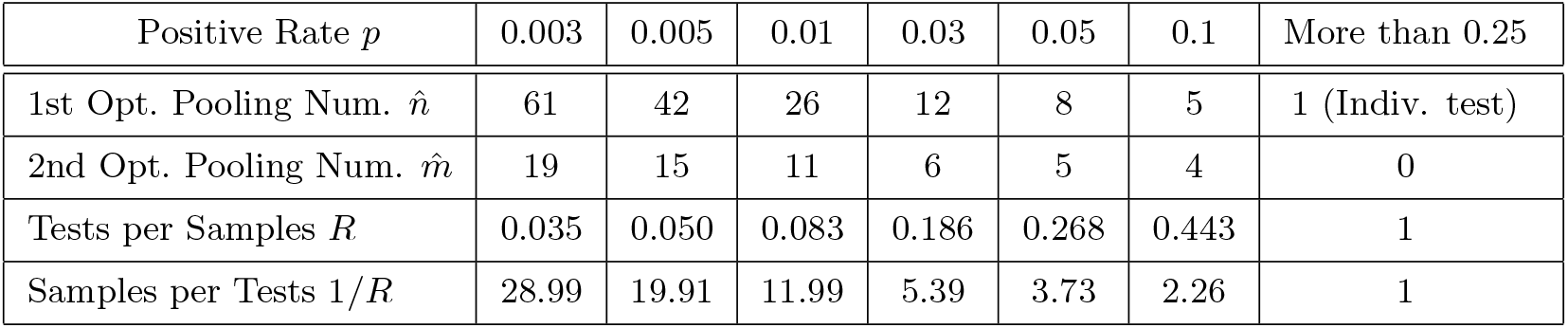
Optimal pooling numbers 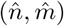 searched and minimum efficiency measure 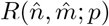 achieved for positive rate *p*

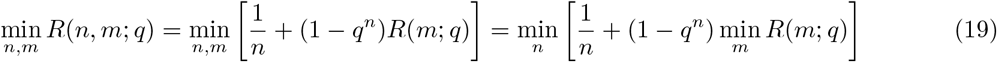

By optimizing the pooling number *n* in the first stage with balancing the minimized efficiency 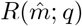 in the second stage, which was 1 due to individual test in the one-stage pool method, a considerable improvement in efficiency is achieved as a whole. As the positive rate *p* becomes smaller, 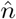 becomes much larger (see Table 1 for comparison). This means that the efficiency can be further improved by excluding negative samples by the pool test using a larger 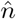 as a *pre-screening* before the conventional one-stage pool test is performed for 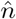 samples in the second stage.

In Table 4, when *p* is large (for example, 0.05, 0.1), the pool test in the second stage is performed with 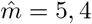 which is close to the pool test in the first stage 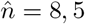. At first glance, it seems that the second stage pool test is useless and the individual test is enough, but the result is still more efficient.

### 3.2 First-order approximation of the optimal solution

The searching of optimal 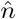 and 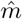 in the two-stage pool test is tedious due to 2-dimensional search, which seems the reason for heuristic and independent selection of *m* in the second stage.

Similarly to the case of one-stage pool method, we consider to expand the exponential terms to the McLaughlin series and approximate them as linear terms by using Bernoulli’s inequality.

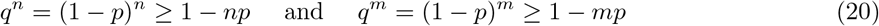

Substituting these into Eq. (18) gives the following equation, the upper bound 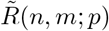.

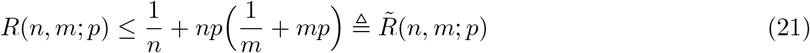

In order to minimize the upper bound 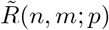, it is partially differentiated with *n* and *m* to zero.

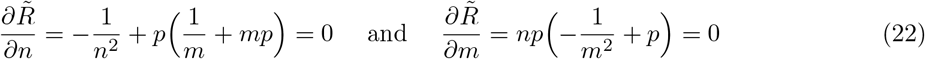

Hence, from these equations, estimated approximate solutions of the optimal numbers 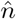 and 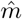 are obtained explicitly as follows.

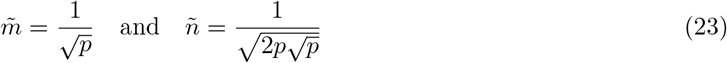

By substituting these into Eq. (21), the minimum value 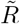 as the upper bound of the minimum value of *R* achieved can be obtained by the following equation, similarly to Eq. (15).

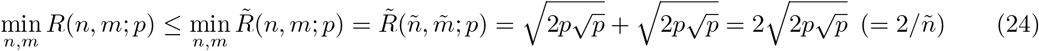

Here, as a condition that satisfies 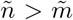 from the subject, we obtain the following condition of *p* from Eq. (23), which corresponds to the limit value *p*^*∗*^ in the one-stage pool test as an approximation.

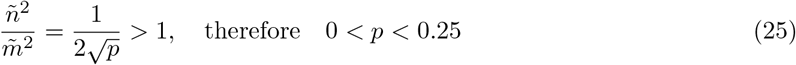

Then, it results in the following.

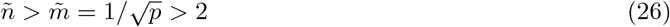

**Table 5** shows the explicit approximate solutions obtained corresponding to Table 4. It can be seen that the smaller the *p*, the tighter the approximation.

However, with the pool method, even though the number of samples *N* direction can be parallelized, if the number of stages increases, the test will wait for the result. Therefore, the maximum inspection time will be longer. For example, if it takes one day for the test result, the normal one-stage pool method takes two days, and the two-stage pool method here takes three days. In addition, there is a concern that performing a pool test with a large number for pooling may dilute the sample and increase the probability of false negatives. In that sense, this two-stage pool method is theoretically highly efficient, but in actual operation it seems that it will be operated with moderate numbers for pooling and no more multi-stages.

**Table 5.** Approximate solutions of optimal 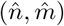 and minimum 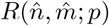 for positive rate *p*: 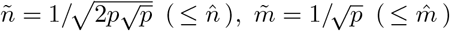, and 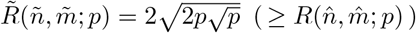 Properly speaking, it states that the estimated optimal pooling numbers are ⌈*ñ*⌉ and 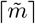 and thereby the optimal efficiency measure less (better) than 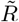 will be achieved.

| Positive Rate $p$ | 0.003 | 0.005 | 0.01 | 0.03 | 0.05 | 0.1 | More than 0.25 |
| --- | --- | --- | --- | --- | --- | --- | --- |
| 1st Opt. Pooling Num. $\tilde{n}$ | 55.16 | 37.61 | 22.36 | 9.81 | 6.69 | 3.98 | 1 (Indiv. test) |
| 2nd Opt. Pooling Num. $\tilde{m}$ | 18.26 | 14.14 | 10.00 | 5.77 | 4.47 | 3.16 | 0 |
| Tests per Samples $\tilde{R}$ | 0.036 | 0.053 | 0.089 | 0.204 | 0.299 | 0.503 | 1 |
| Samples per Tests $1/\tilde{R}$ | 27.58 | 18.80 | 11.18 | 4.91 | 3.34 | 1.99 | 1 |

### 3.3 Estimation of positive rate *p* and adaptive optimization

Unlike the conventional one-stage pool method, in the case of this two-stage pool method, it is difficult to explicitly solve the positive rate *p* from the (average) value 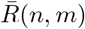 obtained for pre-selected (*n, m*) without knowing *p*. Because, it becomes a higher-order equation of *q* (or *p*) as can be seen from Eq. (18).

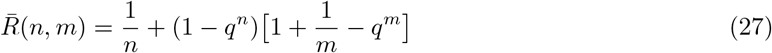

However, if the (average) efficiency measure in the second stage 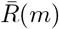 is also available, then we have

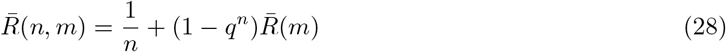

and thereby the exact estimation (solution) of *p* becomes possible as its inverse problem. Actually by solving Eq. (28) for *p* = 1 − *q*, we obtain the following formula.

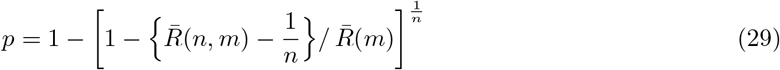

Otherwise, if 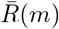 is not available or tedious to further calculate and memorize, then we have to consider some method to estimate the value of *p* from 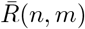 alone.

It is possible from Eq. (21) which approximates the power term in Eq. (27) by Bernoulli’s inequality (McLaughlin expansion). Then, we have the following upper bound on the averaged value 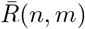.

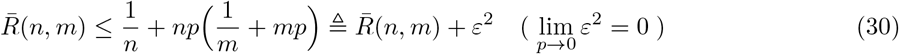

The upper bound on the right side is a quadratic equation of *p*, which is complicated but can be solved and approximated by the following lower bound 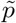.

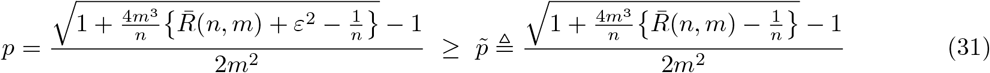

**Table 6** shows the result of the approximate estimation of positive rate *p* obtained in the case of 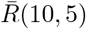 and also the result of the exact estimation using 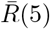 together. It can be seen that the approximate estimation 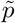 is sufficiently practical including the inequality sign. It is also seen that the smaller the true positive rate *p*, the tighter the approximation 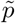, which is preferable in the large scale mass inspection.

Similarly to the case of one-stage pool method, the adaptive optimization scheme in this two-stage pool method could be followed, switching the preset pooling numbers (*n, m*), (10, 5) in Table 6, to the optimal pair via estimating the positive rate *p* or its approximate 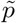 in the above.

**Table 6.** Estimation of positive rate *p* from (average) efficiency measure 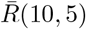 and 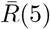 at *n* = 10 and 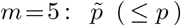 The approximate estimate 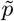 is calculated from 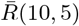 alone by Eq. (31). The exact estimate *p* is calculated by Eq. (29), using also 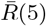 which is already calculated in Table 3.

| Positive Rate $p$ | 0.003 | 0.005 | 0.01 | 0.03 | 0.05 | 0.1 |
| --- | --- | --- | --- | --- | --- | --- |
| Ave. Efficiency $\bar{R}(10, 5)$ | 0.106 | 0.111 | 0.124 | 0.190 | 0.271 | 0.497 |
| Ave. Efficiency $\bar{R}(5)$ | 0.215 | 0.225 | 0.249 | 0.341 | 0.426 | 0.610 |
| Exact Estimate $p$ | 0.003 | 0.005 | 0.010 | 0.030 | 0.050 | 0.100 |
| Approximate $\tilde{p}$ | 0.003 | 0.005 | 0.010 | 0.027 | 0.042 | 0.071 |

## 4. Consideration about false positives and false negatives

It is said that the PCR test is highly sensitive and can detect even a small amount of virus. Therefore, non-infected persons will be negative, but in rare cases, false positives will be obtained due to the adhesion of virus debris or contamination during the test (probability is very low, 1% or less). On the other hand, there is a high probability that the infected person will be false negative (15 to 30%) because the virus was not properly collected in the first place or was destroyed in the processes for PCR.

Although detailed analysis is not performed here, it seems that the pool method does not qualitatively work in the wrong direction and provides a robust framework for those false test results. If it is a false positive, it is tested again at the next individual test, and if it is a false negative, it will not change other positives to false negatives in the sample pool, thus the test results will basically not change in any case and remain the same as the individual test results.

However, as the pooling number *n* becomes larger, especially in the case of two-stage pool test, the probability of false negatives due to reduced test accuracy due to dilution may increase, which seems to be one of the reasons why *n* = 4 or 5 are used in actual pool tests. Thus, if the theoretically optimal pooling number 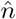 is large corresponding to the case of small positive rate *p*, then the possibility of false negatives due to such dilution should also be taken into account in the actual practice of pool testing.

## 5. Concluding remarks

The method of pooled PCR test has been mathematically formulated in the ordinary one-stage case and also in the multi-stage (here two-stage) case as its natural recursive extension. The efficiency measure *R* defined by the ratio of “tests per samples” is given by a nonlinear function of the pooling number *n* (and *m* in two-stage) and the positive (infection) rate *p* of the testing population, thus the optimal pooling number(s) is difficult to solve together with the difficulty of estimating *p*. Those seem the main reason that the pool tests have been performed in rather intuitive and *ad hoc* manner such as setting *n* = 4 or 5.

In this research, the efficiency measure function *R* is analyzed mathematically in an intensive way, showing the boundary conditions, the efficient range [*A, B*] for *R <* 1, and the limit value *p*\*≐ 0.308 where the pool test becomes inefficient (*R* ≥ 1) beyond that (*p* ≥ *p*^*∗*^) in the case of one-stage pool test.

In addition, by approximating the power term of *R* in a linear form (Bernoulli’s inequality), asymptotic approximate solutions have been explicitly obtained for the optimal pooling numbers 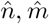, achieved minimum efficiency 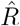, and also for the estimation of positive rate *p*. Thereby, the adaptive optimization scheme of the pool test has been presented. It has been shown that those explicit approximate solutions converge to the true values as the positive rate *p* becomes small, which is preferable in mass inspection.

There are so many related articles and papers on the web [2], actually too many to check and cite each. Thus, some of the analysis here seems to be already known in the research field, however some might be novel. In any case, it will be author’s pleasure if the results here are useful for deeper mathematical insights into and better understanding of the pool inspection, and also in its actual practice.

## Data Availability

no

## Footnotes

1) Actually, the infection curve is expressed by an exponential function with time constant *β − γ*, depending not only on quarantine rate *γ* but also on infection rate *β*. Wearing masks and vaccination are related to reduce *β*.

2) More tight approximate solution will be obtained if we truncate the McLaughlin expansion at the second order of *n* in Eq. (11). However, the case results in a cubic equation of *n*, and it is tedious to solve.

